# Readability Assessment of Patient-Reported Measures Used During Heritable Cancer Genetic Testing

**DOI:** 10.64898/2026.08.13.26360322

**Authors:** Abiodun C Adegbesan, Liesel M FitzGerald, Joanne L Dickinson, Kelsie Raspin, Jessica Roydhouse

**Affiliations:** Menzies Institute for Medical Research, University of Tasmania, Hobart, TAS, 7000, Australia

**Author notes:** These authors contributed equally to this work (Kelsie Raspin, Jessica Roydhouse).

**Keywords:** readability, patient-reported measures, cancer, genetic testing, health literacy

## Abstract

**Background:** Patient-reported measures (PRMs), including patient-reported outcome and experience measures, capture patients’ perspectives on their health status and healthcare experiences. In cancer genetics, PRMs have been used to assess genetic knowledge, psychosocial outcomes, and decision-making. However, patients must understand these measures to provide useful information, an ability which is influenced by general and health literacy levels. Readability guidelines recommend that patient-facing materials be written at or below a Grade 6 level. This study evaluated the readability of PRMs used in a cancer genetic testing context.

**Objective:** To assess whether PRMs used in heritable cancer genetic testing meet recommended readability levels using validated indices.

**Methods:** PRMs were identified from a recent systematic review of PRMs used in heritable cancer genetic testing, which reported 83 instruments across eight categories. English-language PRMs containing structured question items and response scales were eligible for extraction and converted into plain text for analysis. Readability was assessed using four validated indices: Flesch Kincaid Grading Level (FKGL), FORd, CAylor, and STicht (FORCAST) formula, Flesch Reading Ease Score (FRES), and Simple Measure of Gobbledygook (SMOG) via an automated readability software. Descriptive analysis and numerical comparison evaluated readability levels across PRM categories and against the recommended Grade 6 reading level.

**Results:** Sixty-five PRMs met the eligibility criteria, with most, including validated instruments, exceeding the recommended Grade 6 reading level. Across the eight categories, genetics-specific PRMs required the highest readability levels, indicating higher readability demands.

**Conclusions:** Most PRMs, particularly those specific to genetics, do not meet readability guidelines. This may limit their accessibility to individuals with limited general and health literacy. Development of PRMs specific to genetics should consider strategies to improve readability, such as plain-language approaches and involvement of individuals with limited general or health literacy.

## 1 Background

Patient-reported measures (PRMs) encompass both patient-reported experience measures (PREMs) and patient-reported outcome measures (PROMs) (1). PREMs are tools used to capture patients’ perception of their experience with their healthcare and the degree to which their needs were met (2). PROMs capture any report of the status of a patient’s health condition, that comes directly from the patient, without interpretation of the patient’s response by a clinician or anyone else (3). Collectively, they offer essential insights into patients’ experiences of treatment and care, as patients’ perceptions of their symptoms, treatment experiences, and outcomes often differ from those of their clinicians (4).

In oncology, PROMs have been linked to improved patient satisfaction, overall patient survival, fewer hospitalisations, and reduced emergency department visits (5–7). Collection of PREMs has identified systemic issues within oncology services, which help guide quality improvement efforts and policy development to enhance patient satisfaction and outcomes (8). As oncological care increasingly incorporates precision medicine and genetic testing (9), there is growing recognition that PRMs are also important for understanding patient experiences and outcomes in this context (10).

Around 5%–10% of all cancers can be explained by a known germline genetic variant (12). Identifying pathogenic variants through germline genetic testing can inform the selection of appropriate interventions and precision therapies (13). As genetic testing can have psychosocial impacts (e.g., distress) (14), PRMs may provide information that is otherwise unavailable in clinical or administrative data, as evidenced by a 2021 systematic review (15). This review identified PROM use in genetic testing to capture patient-relevant outcomes and concepts including anxiety, knowledge, decision satisfaction, and concerns (15).

Whether PRMs provide useful information largely depends on patients’ ability to adequately read and understand them. This ability is, in turn, determined by the patient’s health literacy (16). Health literacy is defined as the degree to which individuals have the capacity to obtain, process, and understand basic health information and services needed to make appropriate health decisions in the healthcare system (17). Health literacy is built upon general literacy skills such as reading, writing, and speech comprehension (18), which means that individuals with limited general literacy are more likely to experience difficulty when engaging with complex medical information. An individual with low health literacy may be unable to accurately complete health-related surveys and materials, often leading to misunderstanding, incorrect responses, or avoidance, which can compromise their ability to make informed healthcare decisions and affect data quality (19). Similarly, completion of genetics-specific PRMs by those with low health literacy may lead to misinterpretation of medical terminology, inaccurate outcome reporting, and heightened psychological distress, thereby compromising the reliability and validity of the data (20). To address varying literacy levels, patient-facing health materials should be written at or below a Grade 6 reading level (equivalent to a reading age of 11-12 years) (21).

Readability indices are a common tool for evaluating whether patient-facing health materials are written in plain and accessible language (22). Readability is defined as the comprehension level a person must have to understand the written materials and to objectively measure the difficulty of the written materials (23). Readability indices assess the complexity of text by examining features such as sentence length, word length, and syllable count (24). They generate an estimated reading level by indicating the level of education typically required to understand the text with ease. Some of these indices include Flesch Kincaid Grading Level (FKGL) (25), FORd, CAylor, and STicht (FORCAST) (26), Flesch Reading Ease Score (FRES) (27) and Simple Measure of Gobbledygook (SMOG) (28).

Over the past decade, these indices have been used to evaluate the readability levels of PRMs used in various health fields such as ophthalmology (29), sports medicine (30), laryngology (31), head and neck oncology (23), orthopaedics (32) and neurology (headaches) (33). However, there is limited information on the readability of PRMs used in cancer genetics. In this study, we aimed to evaluate the readability of PRMs used in heritable cancer genetic testing identified by a recent systematic review (34).

## 2 Materials and Methods

### 2.1 Identification of PRMs

The selection of PRMs for inclusion in this study was based on a prior systematic review of PRMs (34). In the systematic review, 83 different measures were assessed and classified into eight categories; 1) genetics-specific PRMs (broadly sub-categorised into knowledge and experience, including perceptions, concerns, acceptability), 2) risk-related, 3) health-related, 4) anxiety and depression, 5) worry and distress, 6) decision-making, 7) genetic counselling-related, and 8) others (34) (*Figure 1*).

**Figure 1:**
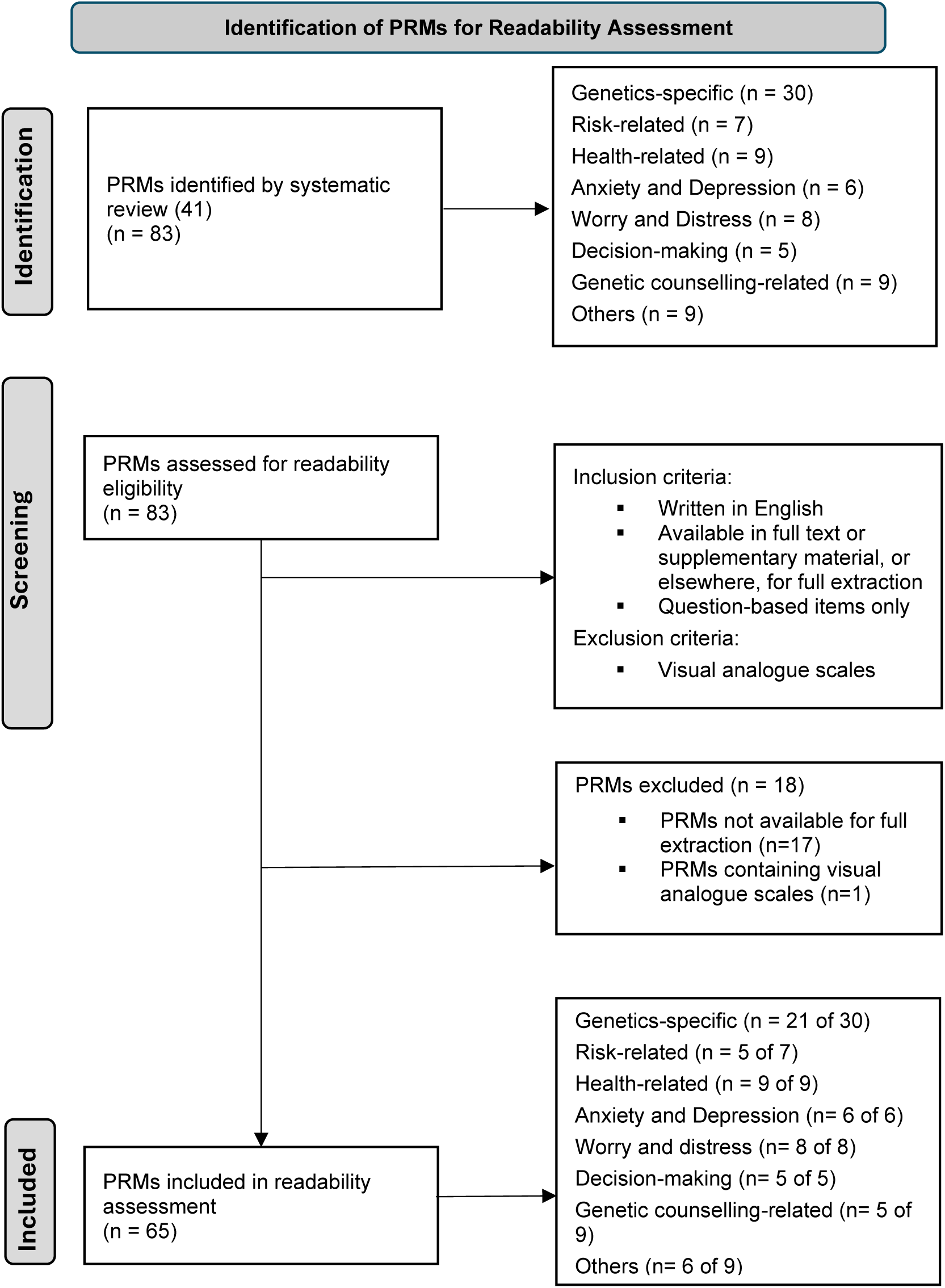
Overview of identification, screening and inclusion of PRMs for readability assessment.

### 2.2 Inclusion Criteria of PRMs

PRMs from the systematic review (34) were included if they were: 1) written in English, 2) available in full text or supplementary material in their entirety, or elsewhere, for full extraction, and 3) structured question-based items (i.e., PRMs using only visual analogue scales were excluded), as shown in *Figure 1*.

### 2.3 Data Preparation of PRMs

All PRMs were extracted from the corresponding studies, transcribed and reformatted into plain-text, prose-style using Microsoft Word. Numbers, bullet points, and layout features were removed to ensure that the readability assessment captured the true linguistic complexity of the instrument, including sentence length and word difficulty, without distortion from formatting elements. This approach aligns with previous readability studies in the literature (35,36), and was necessary because most readability indices (e.g., FORCAST, FRES, SMOG, etc.) are optimised for analysing continuous text (prose format) rather than questionnaire formatting.

As per previous recommendations (35,36), the main questions, the response options, Likert-type scales and instructions were included, if available (31). Copyright practices, disclaimers, acknowledgements, author information, citations and preferences were excluded from the analysis (31).

### 2.4 Readability Indices

The following readability assessment indices were utilised, 1) FORCAST (26), 2) FKGL (25), 3) FRES (27), and 4) SMOG (28). These readability indices were selected due to specific individual strengths and together, provide a comprehensive readability assessment. The use of multiple readability indices has been adopted by many of the readability assessment studies undertaken in the literature, to enable a more nuanced understanding of readability (23,31). FKGL estimates the United States (US) grade level readability of a written material using sentence length and syllable count and is commonly applied in educational and health research (25). FORCAST does not rely on sentence length and focuses on the number of monosyllabic words (26). FRES is widely used for comparing text difficulty and focuses on the average sentence length and syllables per word (27). SMOG is recommended for health and patient education materials and estimates the years of education required to understand a text based on the frequency of polysyllabic words (28). These tools evaluate different linguistic features and together, can provide a comprehensive evaluation of PRM readability.

For SMOG, FKGL, and FORCAST indices, the scores correspond to US Grade levels with lower scores indicating easier readability (i.e., a score of 6 corresponds to a US Grade 6 reading level) (37), with scores ≤ 6-8 considered acceptable readability. In contrast, FRES ranges from 0 – 100, with higher scores indicating easier readability, for example, scores between 60 – 70 correspond roughly to a US Grade 8 reading level, while scores ≥ 80 indicate Grade 6 (37). Here, we report the FRES, in addition to the corresponding grade level for easier comparison with the other indices.

### 2.5 Readability Analysis

We used *readable.com* (38) to conduct our readability analysis. It is an online tool that calculates multiple readability scores equivalent to grade levels and has been used in previous studies (31).

#### Descriptive Analysis

Readability scores for each of the PRMs were calculated across the four readability indices. In addition, each PRM was evaluated against the recommended Grade 6 reading level threshold for each index to determine whether it met the criterion for adequate readability. For all of the readability indices (including the FRES corresponding grade level), lower scores indicated easier readability, while higher scores indicated more difficulty in reading.

#### Comparative Analysis

Readability scores generated using each of the four indices were descriptively compared across the eight PRM categories. Comparative interpretation was based on the overall pattern and distribution of readability scores within each category to identify categories with relatively higher readability burden. We also conducted a numerical comparison of indices across categories to identify which PRM category and sub-category had measures with higher readability demands. As a separate analysis, we examined the readability of validated PRMs developed after the 2009 FDA guidance on PROMs (3). No formal statistical analyses or pooled summary measures were calculated, as analysis was intended to provide a descriptive overview of readability trends across the PRM categories.

## 3 Results

From the 83 PRMs described in our prior systematic review (34), 65 met the inclusion criteria and were assessed for readability *(Figure 1)*. Original references for all PRMs discussed in this section are provided in the Supplementary.

### 3.1 Readability assessment of genetics-specific PRMs

Across the 21 genetics-specific PRMs, readability levels were variable, ranging from Grade 6 to Grade 12, but were generally higher than the recommended Grade 6 reading level (*Tables 1 & 2*). SMOG scores ranged from 7.5 to 15.4, FKGL from 5.8 to 13.7, FRES from 7.0 to 12.0 and FORCAST from 9.3 to 13.3.

Within the genetics-specific PRMs pertaining to genetic knowledge *(Table 1),* five instruments were validated. All these validated PRMs were higher than the recommended Grade 6 reading level across all four readability indices. Similarly, all unvalidated PRMs measuring genetic knowledge (n=3) had reading levels higher than Grade 6 across all four readability indices.

**Table 1:** Readability assessment of genetics-specific PRMs pertaining to genetic knowledge.

| PRM | Year | FKGL<sup>#</sup> | FORCAST<sup>#</sup> | FRES*<br>(corresponding<br>grade level) | SMOG<sup>#</sup> |
| --- | --- | --- | --- | --- | --- |
| *Validated* |  |  |  |  |  |
| Breast Cancer/BRCA Knowledge Scale (1) | 1996 | 6.3 | **10.7** | **70.9 (7.0)** | **7.5** |
| Knowledge of Cancer Genomics (2) | 2014 | **9.4** | **11.4** | **47.3 (12.0)** | **11.8** |
| Breast Cancer Genetic Counselling Knowledge Scale (3) | 2005 | **9.6** | **10.3** | **56.5 (10.0)** | **12.9** |
| National Institute of Health National Centre for Human Genome Research Cancer Genetics Consortium Knowledge Scale (4) | 2004 | **11.3** | **11.4** | **42.2 (12.0)** | **14.2** |
| KnowGene Scale (5) | 2019 | **12.1** | **11.5** | **44.0 (12.0)** | **13.6** |
| *Unvalidated* |  |  |  |  |  |
| Understanding of genetic testing result (6) | 2013 | **9.7** | **12.1** | **47.6 (12.0)** | **12.7** |
| Knowledge of genetic disease (ClinSeq knowledge scale) (7) | 2012 | **10.3** | **11.3** | **50.5 (11.0)** | **13.2** |
| Knowledge of *BRCA* testing and communication with children (8) | 2018 | **11.4** | **12.5** | **35.8 (12.0)** | **13.3** |
| PRMS are ranked in order from the most readable to the least readable (depending on validation status). PRMs above the recommended readability score are in **bold**; <sup>#</sup> Lower scores indicate easier readability; *Higher scores indicate more difficulty reading. SMOG: (Simple Measure of Gobbledygook); FKGL: (Flesch–Kincaid Grade Level); FORCAST (FORd, CAYlor, and StichT); FRES: (Flesch Reading Ease Score); |  |  |  |  |  |

Genetics-specific PRMs pertaining to genetic experience *(Table 2)* had only two validated PRMs (2 of 13); the Multidimensional Impact of Cancer Risk Assessment (MICRA) and the Perceived Personal Control Scale, and both exceeded the recommended Grade 6 readability level across all four readability indices. However, four unvalidated PRMs were found to meet the recommended Grade 6 reading level in the FKGL index. These PRMs are mostly study-specific and included, disclosure of cancer genetic testing results, reasons for declining polygenic risk score testing, cancer-prevention and screening behaviour, and lifestyle changes in response to testing results. Both validated and unvalidated PRMs in this sub-category had FORCAST and SMOG readability scores above Grade 8 except for the Patient Acceptability Scale. The Privacy and Confidentiality Concerns Scale (a study-specific measure), consistently scored above Grade 12 across all readability indices.

**Table 2:** Readability assessment of genetics-specific PRMs pertaining to genetic experience.

| PRM | Year | FKGL<sup>#</sup> | FORCAST<sup>#</sup> | FRES*<br>(corresponding grade level) | SMOG<sup>#</sup> |
| --- | --- | --- | --- | --- | --- |
| *Validated* |  |  |  |  |  |
| Multidimensional Impact of Cancer Risk Assessment (MICRA) (9) | 2002 | **8.5** | **12.7** | **52.9 (12.0)** | **10.4** |
| Perceived Personal Control Scale (10) | 1999 | **7.5** | **9.5** | **67.4 (8.0)** | **11.6** |
| *Unvalidated* |  |  |  |  |  |
| Disclosure of genetic testing results (11) | 2008 | 6.2 | **9.3** | **70.5 (7.0)** | **10.4** |
| Reasons for declining polygenic risk score testing (12) | 2008 | 6.5 | **10.2** | **64.4 (8.0)** | **10.4** |
| Cancer prevention and screening behaviour (13) | 2011 | 5.8 | **10.0** | **66.7 (8.0)** | **11.2** |
| Lifestyle changes in response to testing results (14) | 2021 | 6.5 | **10.7** | **62.0 (9.0)** | **10.5** |
| Process acceptability (15) | 2019 | **7.9** | **10.3** | **62.7 (9.0)** | **11.3** |
| Feelings and reactions to testing results (16) | 2021 | **7.2** | **12.0** | **56.6 (11.0)** | **10.2** |
| Patient Acceptability Scale (17) | 2020 | **11.2** | **10.4** | **62.4 (9.0)** | **8.3** |
| Perception of process of results being returned, feelings about results, feelings about sharing results (18) | 2018 | **8.4** | **12.2** | **53.8 (11.0)** | **11.2** |
| Experience and understanding of genetic testing (19) | 2013 | **9.7** | **12.1** | **47.4 (12.0)** | **12.7** |
| Overall acceptability (20) | 2023 | **11.4** | **13.0** | **41.5 (12.0)** | **15.2** |
| Privacy and confidentiality concerns (16) | 2020 | **13.7** | **13.3** | **13.0 (12.0)** | **15.4** |
| PRMs are ranked in order from the most readable to the least readable (depending on validation status). PRMs above the recommended readability score are in **bold**; <sup>#</sup> Lower scores indicate easier readability; *Higher scores indicate more difficulty reading. SMOG: (Simple Measure of Gobbledygook); FKGL: (Flesch–Kincaid Grade Level); FORCAST (FORd, CAylor, and StichT); FRES: (Flesch Reading Ease Score); |  |  |  |  |  |

### 3.2 Readability assessment of genetic counselling-related PRMs

All genetic counselling-related PRMs *(Table 3)*, despite all being validated (5 of 5), showed consistently high readability levels. Overall these instruments exceeded the recommended Grade 6 readability score across all indices, with SMOG scores consistently above Grade 10 reading levels. Similarly, Genetic Counselling Satisfaction Scale (GCSS) consistently scored Grade 12 and above across all readability indices.

**Table 3:** Readability assessment of genetic counselling-related PRMs.

| PRM | Year | FKGL<sup>#</sup> | FORCAST<sup>#</sup> | FRES*<br>(corresponding<br>grade level) | SMOG<sup>#</sup> |
| --- | --- | --- | --- | --- | --- |
| *Validated* |  |  |  |  |  |
| Satisfaction with Genetic Counselling Scale (21) | 1990 | **7.2** | **9.8** | **67.5 (8.0)** | **10.4** |
| Quality of Care Through the Patient's Eye – GENetic counselling for hereditary Cancer (QUOTE-GENECA) (22) | 2005 | **7.8** | **11.1** | **51.5 (12.0)** | **11.1** |
| Genetic Counselling Outcome Scale (GCOS-24) (23) | 2011 | **7.3** | **9.9** | **67.8 (8.0)** | **11.3** |
| Satisfaction with genetic services (24) | 2002 | **8.5** | **10.6** | **53.7 (11.0)** | **12.6** |
| Genetic Counselling Satisfaction Scale (GCSS) (25) | 2004 | **10.6** | **11.5** | **44.5 (12.0)** | **14.7** |
| PRMS are ranked in order from the most readable to the least readable (depending on validation status). PRMs above the recommended readability score are in **bold**; <sup>#</sup> Lower scores indicate easier readability; *Higher scores indicate more difficulty reading. SMOG: (Simple Measure of Gobbledygook); FKGL: (Flesch–Kincaid Grade Level); FORCAST (FORd, CAylor, and Sticht); FRES: (Flesch Reading Ease Score); |  |  |  |  |  |

### 3.3 Readability assessment of risk-related PRMs

Risk-related PRMs *(Table 4)* comprised both validated and unvalidated instruments, with four of the five measures being validated. Among the validated instruments, the Numeracy Scale had the recommended Grade 6 readability score across the FKGL and FRES indices. In contrast, although validated, the eHealth Literacy Scale (eHEALS) was the least readable PRM within this category, with readability scores above Grade 8 across all indices. Perception of cancer risk(s) was the only unvalidated PRM in this category and exceeded the recommended Grade 6 readability level across all indices with the exception of FKGL.

**Table 4:** Readability assessment of risk-related PRMs.

| PRM | Year | FKGL<sup>#</sup> | FORCAST<sup>#</sup> | FRES*<br>(corresponding<br>grade level). | SMOG<sup>#</sup> |
| --- | --- | --- | --- | --- | --- |
| *Validated* |  |  |  |  |  |
| Numeracy scale (26) | 1997 | 4.0 | **8.5** | **86.2 (6.0)** | **7.6** |
| Subjective Numeracy Scale (27) | 2007 | 6.2 | **9.5** | **73.6 (7.0)** | **9.4** |
| Three-item screening<br>questionnaire for health literacy<br>(28) | 2004 | 8.1 | **9.9** | **62.1 (8.0)** | **11.6** |
| eHealth Literacy Scale<br>(eHEALS) (29) | 2006 | **8.4** | **10.2** | **59.1 (10.0)** | **12.7** |
| *Unvalidated* |  |  |  |  |  |
| Perception of cancer risk(s) (30) | 2017 | 6.9 | **10.7** | **62.4 (8.0)** | **10.4** |
| PRMS are ranked in order from the most readable to the least readable (depending on validation status). PRMs above the recommended readability score are in **bold**; <sup>#</sup> Lower scores indicate easier readability; *Higher scores indicate more difficulty reading. SMOG: (Simple Measure of Gobbledygook); FKGL: (Flesch–Kincaid Grade Level); FORCAST (FORd, CAylor, and StichT); FRES: (Flesch Reading Ease Score); |  |  |  |  |  |

### 3.4 Readability assessment of health-related PRMs

Health-related PRMs *(Table 5)* displayed mixed readability levels despite them all being validated (9 of 9). Generic symptom measures such as the SF-36, SF-12, Brief Symptom Inventory (BSI), Symptom Checklist 90-Revised (SCL-90-R) and the cancer-specific EORTC QLQ-C30 were generally more readable with recommended FKGL scores of Grades 4–6. In contrast, symptom-specific measures, including Sexual Health Inventory for Men (SHIM) and the AUA Symptom Scale had readability scores of Grade 10 and above across all indices.

**Table 5:** Readability assessment of health-related PRMs.

| PRM | Year | FKGL<sup>#</sup> | FORCAST<sup>#</sup> | FRES*<br>(corresponding<br>grade level) | SMOG<sup>#</sup> |
| --- | --- | --- | --- | --- | --- |
| *Validated* |  |  |  |  |  |
| Brief Symptom Inventory (BSI) (31) | 1983 | 4.6 | **10.6** | **75.7 (7.0)** | **7.2** |
| Symptom Checklist-90-Revised (SCL-90-R®) (32) | 1973 | 5.2 | **11.4** | **70.0 (8.0)** | **7.5** |
| EORTC Quality of Life Questionnaire – Core Questionnaire (EORTC-QLQ-C30) (33) | 1995 | 5.7 | **10.1** | **68.9 (8.0)** | **9.4** |
| SF-12 Health Survey (SF-12) (34) | 1994 | 6.3 | **9.0** | **73.7 (7.0)** | **10.0** |
| SF-36 Health Survey (SF-36) (35) | 1992 | 6.1 | **9.8** | **68.9 (8.0)** | **9.8** |
| Gastrointestinal scale from the Giessen Complaints Inventory (36) | 1995 | **6.6** | **11.5** | **59.2 (10.0)** | **9.1** |
| American Urological Association (AUA) Symptom Scale (37) | 1992 | **10.4** | **10.0** | **54.9 (11.0)** | **13.2** |
| Eastern Cooperative Oncology Group Performance Status (ECOG Performance Status) (38) | 1982 | **8.2** | **11.5** | **52.0 (12.0)** | **13.0** |
| Sexual Health Inventory for Men (SHIM) (or Erectile Function – 5 items; IIEF-5 (39) | 1999 | **10.6** | **11.6** | **47.5 (12.0)** | **14.0** |
| PRMs are ranked in order from the most readable to the least readable (depending on validation status). PRMs above the recommended readability score are in **bold**; <sup>#</sup> Lower scores indicate easier readability; *Higher scores indicate more difficulty reading. SMOG: (Simple Measure of Gobbledygook); FKGL: (Flesch–Kincaid Grade Level); FORCAST (FORd, CAylor, and StichT); FRES: (Flesch Reading Ease Score); |  |  |  |  |  |

### 3.5 Readability assessment of anxiety and depression PRMs

All PRMs assessing psychological domains, anxiety and depression (*Table 6*), were validated (6 of 6). DASS-21, STAI, GAD-7 and PHQ-4 were generally more readable with recommended FKGL scores of Grades 3–6. In contrast, the other two PRMs, MAX-PC and HADS had readability scores above Grade 6 for all four indices.

**Table 6:** Readability assessment of anxiety and depression PRMs.

| PROM | Year | FKGL<sup>#</sup> | FORCAST<sup>#</sup> | FRES*<br>(corresponding<br>grade level) | SMOG<sup>#</sup> |
| --- | --- | --- | --- | --- | --- |
| *Validated* |  |  |  |  |  |
| State-Trait Anxiety Inventory (STAI) (40) | 1983 | 3.6 | **9.6** | **78.4 (7.0)** | **7.6** |
| Patient Health Questionnaire – 4 items (PHQ-4) (41) | 2009 | 4.8 | **10.6** | **74.8 (7.0)** | **8.3** |
| Depression Anxiety Stress Scales Short Form (DASS-21) (42) | 2005 | 5.6 | **9.3** | **75.4 (7.0)** | **9.1** |
| Memorial Anxiety Scale for Prostate Cancer (MAX-PC) (43,44) | 2003 | **7.6** | **10.5** | **68.2 (8.0)** | **9.0** |
| Hospital Anxiety and Depression scale (HADS) (45) | 1983 | **8.4** | **8.8** | **71.2 (7.0)** | **9.9** |
| Generalized Anxiety Disorder – 7 items (GAD-7) (46) | 2006 | 6.0 | **11.4** | **66.5 (8.0)** | **8.8** |
| PRMS are ranked in order from the most readable to the least readable (depending on validation status). PRMs above the recommended readability score are in **bold**; <sup>#</sup> Lower scores indicate easier readability; *Higher scores indicate more difficulty reading. SMOG: (Simple Measure of Gobbledygook); FKGL: (Flesch–Kincaid Grade Level); FORCAST (FORd, CAylor, and StichT); FRES: (Flesch Reading Ease Score); |  |  |  |  |  |

### 3.6 Readability assessment of worry and distress PRMs

Within the worry and distress PRMs category *(Table 7)*, five of the eight instruments were validated. Only three of the five validated PRMs, namely PSS-10, Concerns about Recurrence Questionnaire, and Impacts of Events Scale had recommended FKGL scores. Similarly, three unvalidated PRMs; Impact of Cancer Worry, Frequency of Cancer Worry and Breast and Ovarian Cancer Worry scored recommended FKGL scores. The Distress Thermometer, despite being validated, was the least readable with high FORCAST and FRES scores of Grade 12.

**Table 7:** Readability assessment of worry and distress PRMs.

| PRM | Year | FKGL<sup>#</sup> | FORCAST<sup>#</sup> | FRES*<br>(corresponding grade level) | SMOG<sup>#</sup> |
| --- | --- | --- | --- | --- | --- |
| *Validated* |  |  |  |  |  |
| Perceived Stress Scale – 10-items (PSS-10) (47) | 1983 | 6.0 | **8.4** | 80.2 (6.0) | **8.8** |
| Impact of Events Scale – R (48) | 2007 | 5.0 | **9.9** | **76.5 (7.0)** | **7.9** |
| Concerns about Recurrence Questionnaire (49) | 2015 | 6.1 | **10.0** | **68.4 (8.0)** | **9.0** |
| Cancer Worry Scale (CWS) (50) | 1991 | **7.5** | **11.1** | **61.7 (9.0)** | **9.2** |
| Distress Thermometer (NCCNDT) (51) | 2003 | **7.3** | **12.5** | **50.4 (12.0)** | **8.6** |
| *Unvalidated* |  |  |  |  |  |
| Impact of Cancer Worry (52) | 2018 | 6.1 | **11.3** | **66.4 (8.0)** | **9.4** |
| Breast and ovarian cancer worry items (53) | 2003 | 6.6 | **10.7** | **66.8 (8.0)** | **10.1** |
| Frequency of Cancer Worry (52) | 2018 | 6.4 | **12.8** | **63.0 (9.0)** | **8.8** |
| PRMS are ranked in order from the most readable to the least readable (depending on validation status). PRMs above the recommended readability score are in **bold**; <sup>#</sup> Lower scores indicate easier readability; *Higher scores indicate more difficulty reading. SMOG: (Simple Measure of Gobbledygook); FKGL: (Flesch–Kincaid Grade Level); FORCAST (FORd, CAYlor, and StichT); FRES: (Flesch Reading Ease Score); |  |  |  |  |  |

### 3.7 Readability assessment of decision-making PRMs

All five decision-making PRMs *(Table 8)* were validated but showed variable readability levels. The Decisional Conflict Scale, Decision Self Efficacy Scale, and Decision Regret Scale had recommended FKGL readability scores. In contrast, the Satisfaction with Decision Scale and Control Preferences Scale, reported high SMOG, FRES and FORCAST scores above Grade 10, despite their validated status.

**Table 8:** Readability assessment of decision-making PRMs.

| PRM | Year | FKGL<sup>#</sup> | FORCAST<sup>#</sup> | FRES*<br>(corresponding<br>grade level) | SMOG<sup>#</sup> |
| --- | --- | --- | --- | --- | --- |
| *Validated* |  |  |  |  |  |
| Decision Self Efficacy Scale (DSES) (54) | 2002 | 4.9 | **9.7** | **76.9 (7.0)** | **8.4** |
| Decisional Conflict Scale (DCS) (55) | 1995 | 4.7 | **9.8** | **76.8 (7.0)** | **8.7** |
| Decision Regret Scale (56) | 2003 | 5.6 | **10.2** | **72.3 (7.0)** | **9.5** |
| Control Preferences Scale (57) | 1997 | **8.2** | **11.1** | **55.2 (11.0)** | **12.7** |
| Satisfaction with Decision Scale (SWD) (58) | 1996 | **9.0** | **11.6** | **49.7 (12.0)** | **13.0** |
| PRMS are ranked in order from the most readable to the least readable (depending on validation status). PRMs above the recommended readability score are in **bold**; <sup>#</sup> Lower scores indicate easier readability; *Higher scores indicate more difficulty reading. SMOG: (Simple Measure of Gobbledygook); FKGL: (Flesch–Kincaid Grade Level); FORCAST (FORd, CAylor, and StichT); FRES: (Flesch Reading Ease Score); |  |  |  |  |  |

### 3.8 Readability assessment of other PRMs

Other PRMs *(Table 9),* although predominantly validated (5 out of 6 PRMs), had variable readability. Validated measures, such as the Breast Q Reduction Module, Interpersonal Support Evaluation List (ISEL) and Parents-Adolescent Comunication Scale (PACS), achieved recommended FKGL readability scores. In contrast, the other two PRMs, Threatening Medical Situation Inventory (TMSI) and Telemedicine Satisfaction Questionnaire had readability scores above Grade 6 for all four indices. The perception and experience PRM, an unvalidated study-specific measure, was the least readable in this category consistently scoring above Grade 10 across all indices.

**Table 9:**
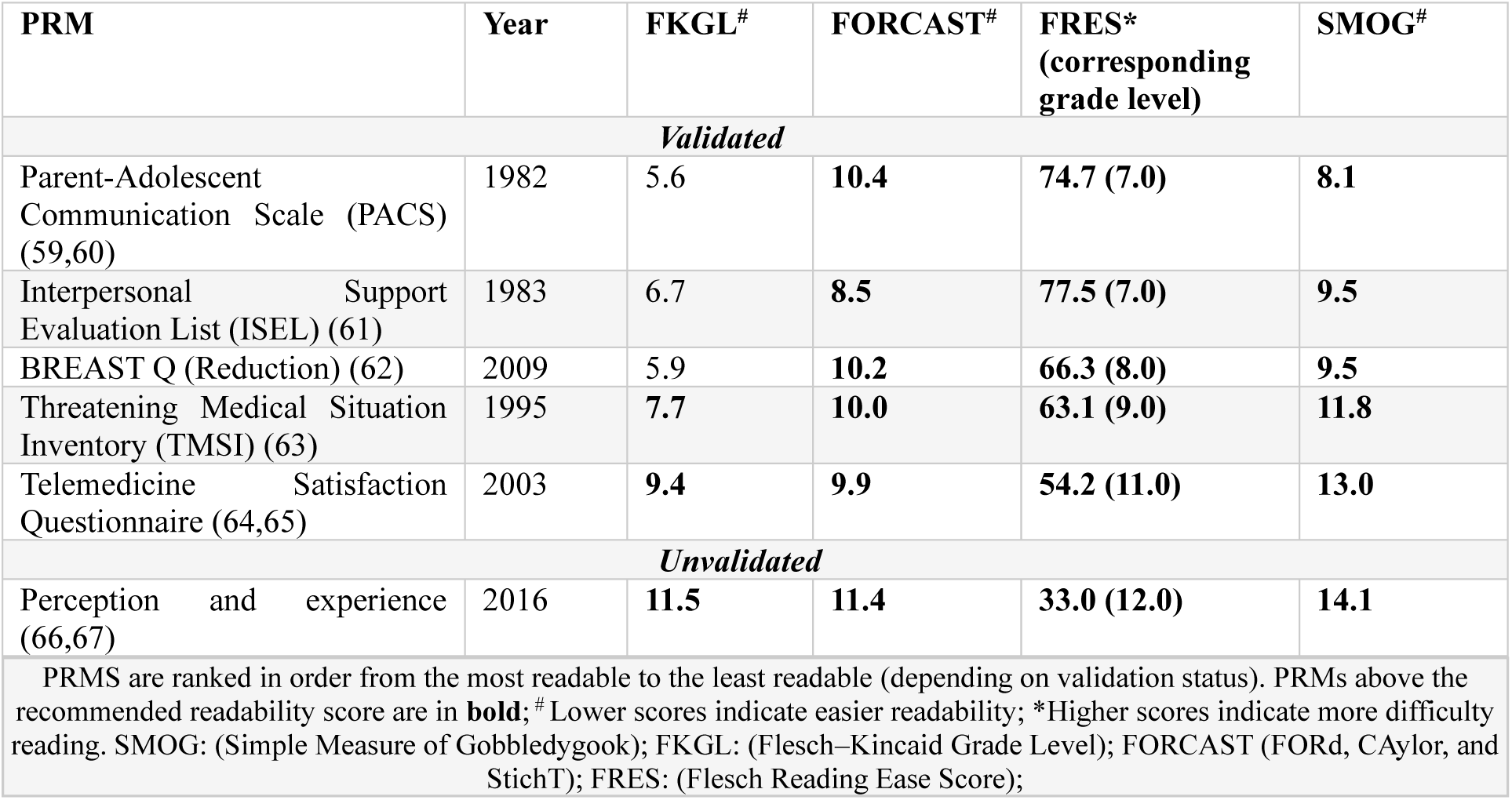
Readability assessment of other PRMs.

### 3.9 Validated PRMs developed after the FDA’s 2009 guidance document

Six validated PRMs developed after the FDA’s 2009 guidance document were identified across five categories *(Supplementary Table 1)*, namely, 1) genetics-specific (Knowledge of Cancer Genomics (2014) and the Knowgene Scale (2019)), 2) genetic counselling-related (Genetic Counselling Outcome Scale (GCOS-24) (2011)), 3) worry and distress (Concerns about Recurrence Questionnaire (2015)), 4) anxiety and depression (Patient Health Questionnaire (PHQ-4) (2009)) and 5) other (Breast Q (reduction) (2009)). All these PRMs were above the recommended readability levels, with SMOG and FORCAST scores above the Grade 6 reading level. Knowledge of Cancer Genomics and the Knowgene Scale, both measuring genetic knowledge, were the least readable PRMs after 2009, with readability scores mostly above Grade 10 across all readability indices.

## 4 Discussion

Our study is the first to assess the readability of PRMs used in the context of heritable cancer genetic testing. Notably, most PRMs exceeded the recommended Grade 6 reading level for health-related information, which was evident across all PRM categories, though the PRMs specific to genetics demonstrated the highest reading complexity. Most PRMs within the genetics-specific category, including knowledge and experience measures, demonstrated higher readability scores relative to PRMs in other categories, with readability scores predominantly above the Grade 10 reading level. Both validated and unvalidated PRMs tended to impose greater readability demands across all the PRM categories, and nearly all PRMs developed after the 2009 FDA guidance had readability scores above the recommended grade levels.

Our findings are consistent with previous studies demonstrating increased readability demands in PRMs, specifically PROMs, across various oncological and non-oncological settings. For example, Papadakos *et al.,* (2019) (42) and Uppal *et al.,* (2025) (43), in their readability assessments of PROMs used in the clinical cancer setting and surgical oncology, respectively, reported that more than 70% of the identified measures exceeded the recommended Grade 6 reading level. Our analyses also found that several widely used and validated PRMs, including some of those in the health-related category exceeded the recommended Grade 6 reading level across multiple readability indices, which is consistent with other studies (33,42,44). Most notably, our findings suggest that genetics-specific tools in particular may pose challenges to individuals with lower general or health literacy. This is an important issue to address given the growing importance of patient-centred care in oncology, including the integration of genetic testing into routine cancer care.

Since many PRMs specific to genetics were study-specific, development of future PRMs specific to genetics should incorporate established health literacy and plain language guidelines during item and instruction development, such as those outlined by the American Medical Association and the Agency for Healthcare Research and Quality (40,45). Furthermore, involving patients in the development of these PRM instruments is one potential approach for improving readability. This is particularly important in the context of cancer genetic testing, where complex scientific information must be communicated clearly. Although such involvement is reported in many PROM development studies, the extent and depth of engagement varies considerably, with over one-quarter of studies reporting no patient involvement at any stage of instrument development (46). Concerningly, individuals with lower literacy skills have often been excluded from PROM development processes (47), despite potentially being the least likely to understand the survey items, response sets (48), or complete the PROMs correctly (48). Adopting an inclusive approach to PRM development, with an emphasis on literacy, would be beneficial and generate readable, accessible PRMs across all literacy and cultural gradients.

Although our overall findings were consistent with the literature, the specific scores generated by each readability index differed from those reported in previous studies. One possible explanation is the heterogeneity in methodological approaches for examining readability. In our study, we evaluated PRMs in their entirety, incorporating instructions, survey items, and response options into a single plain text for analysis. By contrast, some prior studies have disaggregated PRM instruments into discrete components such as instructions, individual items and response options, and analysed them independently (49,50). One study reported higher readability scores for the instruction component of the State Trait Anxiety Inventory (STAI) (SMOG 10.3) compared with the survey-item text (SMOG 7.4), suggesting that instructions might be harder to read because they typically contain longer sentences, medical terms and are often excluded from the psychometric validation process (36). When we considered both the instructions and items together for STAI, the SMOG score was 7.6. Although both findings suggest that this item is above the Grade 6 reading level, the variability in inclusion of different PRM components highlights the need for detailed and clear reporting of methodological approaches.

Another source of heterogeneity is the variation in grades produced by different readability indices for the same PRM. We found that readability indices generally identified similar overall patterns of readability difficulty, but the range of the scores differed. This may be due to differences in linguistic features assessed by individual indices (24). For example, our results demonstrated that FKGL scores were consistently of a lower grade level compared to the three other indices. This was in keeping with Freda *et al.,* (2005), who found that the FKGL index often scored patient education materials two to three grade levels lower than SMOG (51). One explanation may be the focus of FKGL on sentence length and syllable count, making it less sensitive to the inclusion of specialised medical terminology. Consequently, FKGL may underestimate the reading difficulty of health-related materials, particularly when texts contain short sentences but complex vocabulary (52), which may be the case for genetics-specific PRMs. Though prior studies have employed only one or two readability indices (22,43), our study suggests that the use of complementary readability indices may provide a more comprehensive picture of readability as they evaluate different aspects of textual complexity.

The choice of software for readability analyses may also produce variation between studies. Our study utilised the widely used *readable.com* software (38), while other studies have used the Oleander Readability Studio Professional Edition (53) platform, and others have performed the analysis manually (42). These methodological variations may partly explain the discrepancies in reported readability scores for the same instrument across studies even when using the same readability indices. For example, in our study, the American Urological Association (AUA) Symptom Scale (54) had a SMOG score of 13.2, compared to 11.2 reported by Hu *et al.,* (2024) (54), who used the Oleander Readability Studio Professional Edition. One possible explanation for variation in readability scores across software is differences in how each tool handles text preprocessing, syllable counting, and proprietary adjustments (38,53). This is in keeping with findings by Mac and colleagues (2023) (55), who suggest that automated readability scores are inconsistent and often inaccurate. Our findings and the heterogeneity in the literature support the Mac and colleagues’ recommendation for the development of comprehensive guidance on the conduct and reporting of readability assessments (55).

Our study had several limitations. Firstly, although some readability indices used in this study, such as the SMOG formula, have demonstrated cross-linguistic validity (i.e., English, French and Spanish), our readability analysis was restricted to the English versions of the included PRMs. Therefore, our study findings may not be generalisable to non-English versions of these measures. Secondly, commonly used readability indices assess reading difficulty of a text using features such as average sentence length and syllable density, however, they do not account for other factors that may influence patient readability and comprehensibility, such as text layout, font size or type, use of visual aids and complex medical terminologies (56). Finally, only 65 of the 83 previously identified PRMs (34) were eligible for readability assessment in this present study due to our inability to find full text versions. Therefore, our findings may not necessarily generalise to the measures that were excluded.

## 5 Conclusion

Most PRMs used in heritable cancer genetic testing, particularly those that are genetics-specific, exceed the recommended Grade 6 reading level, indicating an accessibility gap. These findings underscore the urgent need to develop genetics-specific PRMs that are clear, readable and accessible to patients with lower general or health literacy. Addressing this gap is increasingly critical as patient-centred cancer care, including the integration of genomics into clinical practice, continues to grow.

## Abbreviations

PRMs: Patient-reported measures
PROMs: Patient-reported outcome measures
PREMs: Patient-reported experience measures
FKGL: Flesch Kincaid Grading Level
FRES: Flesch Reading Ease Score
FORCAST: FORd, CAylor, and StichT
SMOG: Simple Measure of Gobbledygook
FDA: Food and Drug Administration

## AUTHOR CONTRIBUTIONS

## FUNDING

DECLARATION OF INTERESTS

## Data Availability

All data produced in the present work are contained in the manuscript

https://doi.org/10.1007/s40271-025-00784-0

## Notes

### Competing Interest Statement

The authors have declared no competing interest.

